# Head-to-head comparison of SARS-CoV-2 antigen-detecting rapid test with professional-collected nasal versus nasopharyngeal swab

**DOI:** 10.1101/2020.12.03.20243725

**Authors:** Andreas K. Lindner, Olga Nikolai, Chiara Rohardt, Susen Burock, Claudia Hülso, Alisa Bölke, Maximilian Gertler, Lisa J. Krüger, Mary Gaeddert, Frank Tobian, Federica Lainati, Joachim Seybold, Terry C. Jones, Jörg Hofmann, Jilian A. Sacks, Frank P. Mockenhaupt, Claudia M. Denkinger

**Author notes:** Correspondence: Claudia M. Denkinger, Division of Clinical Tropical Medicine, Heidelberg University Hospital, Im Neuenheimer Feld 672, 69120 Heidelberg, Germany. Authors contributed equally (Andreas K. Lindner, Olga Nikolai, Frank P. Mockenhaupt, Claudia M. Denkinger).

## Abstract

**Background:** Nasopharyngeal (NP) swab samples for antigen-detecting rapid diagnostic tests (Ag-RDTs) require qualified healthcare professionals and are frequently perceived as uncomfortable by patients.

**Methods:** We performed a manufacturer-independent, prospective diagnostic accuracy study, comparing professional-collected nasal mid-turbinate (NMT) to nasopharyngeal swab, using the test kits of a WHO-listed SARS-CoV-2 Ag-RDT (STANDARD Q COVID-19 Ag Test, SD Biosensor), which is also being distributed by Roche. Individuals with high suspicion for COVID-19 infection were tested. The reference standard was RT-PCR using a combined oro-/nasopharyngeal swab sample. Percent positive and negative agreement, as well as sensitivity and specificity were calculated.

**Results:** Among the 179 participants, 41 (22.9%) tested positive for SARS-CoV-2 by RT-PCR. The positive percent agreement of the two different sampling techniques for the Ag-RDT was 93.5% (CI 79.3-98.2). The negative percent agreement was 95.9% (CI 91.4-98.1). The Ag-RDT with NMT-sampling showed a sensitivity of 80.5% (33/41 PCR positives detected; CI 66.0-89.8) and specificity of 98.6% (CI 94.9-99.6) compared to RT-PCR. The sensitivity with NP-sampling was 73.2% (30/41 PCR positives detected; CI 58.1-84.3) and specificity was 99.3% (CI 96.0-100). In patients with high viral load (>7.0 log_10_ SARS-CoV-2 RNA copies/swab), the sensitivity of the Ag-RDT with NMT-sampling was 100% and 94.7% with NP-sampling.

**Conclusion:** This study demonstrates that sensitivity of a WHO-listed SARS-CoV-2 Ag-RDT using a professional nasal-sampling kit is at least equal to that of the NP-sampling kit, although confidence intervals overlap. Of note, differences in the IFUs of the test procedures could have contributed to different sensitivities. NMT-sampling can be performed with less training, reduces patient discomfort, and it enables scaling of antigen testing strategies. Additional studies of patient self-sampling should be considered to further facilitate the scaling-up of Ag-RDT testing.

## To the Editor

Antigen-detecting rapid diagnostic tests (Ag-RDTs) are likely to play a substantial role in innovative testing strategies for SARS-CoV-2 [1, 2]. Currently, most Ag-RDTs require nasopharyngeal (NP) sampling performed by qualified healthcare professionals. Nasal sampling would enable scaling of antigen testing strategies. The term nasal sampling is often not used uniformly but can be differentiated in anterior nasal sampling (entire absorbent tip of the swab, usually 1 to 1.5 cm, inserted into nostril), and nasal mid-turbinate (as described below) [3].

We conducted a prospective diagnostic accuracy study with the objective to directly compare the performance of professional-collected nasal mid-turbinate (NMT) versus NP swab, using a WHO-listed SARS-CoV-2 Ag-RDT. The reference standard was RT-PCR collected from a combined NP/oropharyngeal (OP) swab. The study was continued until 30 positive NP swab samples according to Ag-RDT were obtained, which is the minimum recommended by the WHO Emergency Use Listing Procedure to demonstrate sample type equivalency [4]. This manufacturer-independent study was conducted in partnership with the Foundation of Innovative New Diagnostics (FIND), the WHO collaborating centre for COVID-19 diagnostics.

Adults at high risk for SARS-CoV-2 infection according to clinical suspicion who attended the ambulatory SARS-CoV-2 testing facility of Charité University Hospital Berlin, Germany, were enrolled from 11-18 November 2020. Participants were excluded if either of the swabs for the Ag-RDT or the RT-PCR reference standard could not be collected.

Participants had to blow once the nose with a tissue. Afterwards, a NMT-sample was collected on both sides of the nose, using the specific nasal swab provided in the test kit of the manufacturer, according to the instructions for use, which also correspond to the U.S. CDC instructions [3]. Briefly, while tilting the patient’s head back 70 degrees, the swab was inserted about 2cm into each nostril, parallel to the palate until resistance was met at turbinates, then rotated 3-4 times against the nasal walls. Subsequently, a separate NP-swab (provided in the manufacturer test kit) for the Ag-RDT and a combined OP/NP-swab (eSwab from Copan placed in 1ml Amies medium) as per institutional recommendations for RT-PCR were taken from different sides of the nose.

The Ag-RDT evaluated was the STANDARD Q COVID-19 Ag Test (SD Biosensor, Inc. Gyeonggi-do, Korea; henceforth called STANDARD Q) [5]. Study procedures followed the same process as described in the prior study by Lindner et al [6]. While the test is commercially available as NP-sampling kit, the nasal-sampling kit is currently available for ‘research use only’ by the manufacturer. The instructions for use of the two test kits showed differences, with a more elaborate extraction process (stirring the swab at least 10 vs. 5 times) and a higher volume of extracted specimen (4 vs. 3 drops) used for testing of nasal samples.

Of 181 patients invited, 180 (99.4%) consented to participate. One patient was excluded as both swabs for the Ag-RDT could not be obtained. The average age of participants was 36.2 years (Standard Deviation [SD] 12.2) with 48.0% female and 14.5% having comorbidities. On the day of testing, 96.1% of participants had one or more symptoms consistent with COVID-19. Duration of symptoms at the time of presentation on average was 4.2 days (SD 2.6). Among the 179 participants, 41 (22.9%) tested positive for SARS-CoV-2 by RT-PCR (Table 1).

**TABLE 1.** Antigen-detecting RDT results with a professional-collected NMT swab and NP swab in RT-PCR positive patients from combined OP/NP swab. CT-values and viral load (in descending order) of the paired RT-PCR samples are shown, as well as the duration of symptoms per patient. The positive percent agreement between NMT and NP samples on Ag-RDT, and the respective sensitivities compared to RT-PCR are shown.

| No. | NMT swab<br>SD Q Ag-RDT | NP swab<br>SD Q Ag-RDT | OP/NP swab RT-PCR |  | Symptom<br>duration (days) |
| --- | --- | --- | --- | --- | --- |
|  |  |  | CT value | Viral load<sup>3</sup> |  |
| 1 | pos (+++) | pos (+++) | 16.41<sup>2</sup> | 9.11 | - |
| 2 | pos (++) | pos (+++) | 19.25<sup>1,4</sup> | - | 7 |
| 3 | pos (+++) | pos (+++) | 19.82<sup>1</sup> | 8.85 | 4 |
| 4 | pos (++) | pos (+++) | 17.68<sup>2</sup> | 8.73 | 1 |
| 5 | pos (+++) | pos (+++) | 17.82<sup>2</sup> | 8.69 | 3 |
| 6 | pos (+++) | pos (+++) | 18.57<sup>2</sup> | 8.46 | 4 |
| 7 | pos (+++) | pos (+++) | 22.09<sup>1</sup> | 8.17 | 4 |
| 8 | pos (++) | pos (+++) | 19.96<sup>2</sup> | 8.05 | 8 |
| 9 | pos (++) | pos (+++) | 22.70<sup>1</sup> | 7.99 | 5 |
| 10 | pos (++) | pos (++) | 20.74<sup>2</sup> | 7.82 | 3 |
| 11 | pos (+++) | pos (+++) | 23.54<sup>1</sup> | 7.75 | 3 |
| 12 | pos (+++) | pos (+++) | 24.09<sup>1</sup> | 7.58 | 3 |
| 13 | pos (+++) | pos (+++) | 24.37<sup>1</sup> | 7.50 | 2 |
| 14 | pos (+++) | pos (++) | 21.83<sup>2</sup> | 7.50 | 5 |
| 15 | pos (+) | pos (+++) | 24.41<sup>1</sup> | 7.49 | 2 |
| 16 | pos (++) | pos (+++) | 24.62<sup>1</sup> | 7.43 | 6 |
| 17 | pos (+) | neg | 22.42<sup>2</sup> | 7.32 | 10 |
| 18 | pos (+++) | pos (+++) | 25.38<sup>1</sup> | 7.20 | 4 |
| 19 | pos (+++) | pos (++) | 22.85<sup>2</sup> | 7.19 | 5 |
| 20 | pos (++) | pos (+++) | 23.05<sup>2</sup> | 7.13 | 5 |
| 21 | pos (++) | neg | 26.48<sup>1</sup> | 6.87 | 8 |
| 22 | pos (+++) | pos (++) | 24.13<sup>2</sup> | 6.81 | 3 |
| 23 | pos (+++) | pos (+++) | 24.63<sup>2</sup> | 6.66 | 2 |
| 24 | pos (+++) | pos (++) | 24.71<sup>2</sup> | 6.64 | 3 |
| 25 | pos (+++) | pos (++) | 27.84<sup>1</sup> | 6.47 | 2 |
| 26 | neg | neg | 25.55<sup>2</sup> | 6.39 | 4 |
| 27 | pos (++) | pos (+++) | 28.82<sup>1</sup> | 6.18 | 5 |
| 28 | pos (+++) | pos (++) | 26.64<sup>2</sup> | 6.07 | 3 |
| 29 | neg | neg | 26.79<sup>2</sup> | 6.02 | 3 |
| 30 | pos (+++) | pos (++) | 29.87<sup>1</sup> | 5.87 | 4 |
| 31 | neg | neg | 30.91<sup>1</sup> | 5.56 | 7 |
| 32 | pos (+++) | pos (+++) | 29.31<sup>2</sup> | 5.27 | 5 |
| 33 | neg | neg | 29.84<sup>2</sup> | 5.12 | 4 |
| 34 | pos (+) | neg | 33.05<sup>1</sup> | 4.93 | 8 |
| 35 | pos (++) | pos (+) | 31.56<sup>2</sup> | 4.61 | 5 |
| 36 | neg | neg | 34.24<sup>1</sup> | 4.58 | 6 |
| 37 | neg | neg | 34.52<sup>1</sup> | 4.50 | 9 |
| 38 | pos (+) | pos (++) | 34.69<sup>1</sup> | 4.44 | - |
| 39 | pos (+) | neg | 35.16<sup>1</sup> | 4.31 | 10 |
| 40 | neg | neg | 34.58<sup>2</sup> | 3.71 | 7 |
| 41 | neg | pos (+) | 35.53<sup>2</sup> | 3.43 | 8 |
|  | Sensitivity<br>33/41 (80.5%) | Sensitivity<br>30/41 (73.2%) | <sup>1</sup> Roche Cobas SARS-CoV-2 assay (E-gene, T2 target)<br><sup>2</sup> TibMolbiol assay, E-gene target.<br><sup>3</sup> log<sub>10</sub> SARS-CoV-2 RNA copies/swab<br><sup>4</sup> only T1 CT value, T2 invalid due to suspected mutation within oligonucleotide binding region. VL not specified. |  |  |
|  | Positive percent agreement<sup>5</sup><br>93.5% (CI 79.3-98.2) |  |  |  |  |

**Abbreviations:** No., patient number; SD Q, STANDARD Q COVID-19 Ag Test (SD Biosensor); Ag-RDT, antigen-detecting rapid diagnostic test; NMT, nasal mid-turbinate; NP, nasopharyngeal; OP, oropharyngeal; CT, cycle threshold; RT-PCR, reverse transcription-polymerase chain reaction; neg., negative; pos (+), weak positive; pos. (++) , positive; pos. (+++) , strong positive.

No invalid Ag-RDT results were observed on either NMT- or NP-samples. Four patients tested positive by NMT- but not by NP-sampling. One patient was positive by NP-sampling only. The positive percent agreement was 93.5% (95% CI 79.3-98.2); including one false positive result with NMT and one with NP. The negative percent agreement was 95.9% (95% CI 91.4-98.1). Inter-rater reliability was high (kappa 0.95 for NMT; 0.98 for NP). In the semi-quantitative read-out of the test band intensity in double positive pairs, there was no remarkable difference (8 higher on NMT, 9 higher on NP). A third reader was necessary for the agreement on the results of three tests for which the test band was very weak.

The STANDARD Q Ag-RDT with NMT-sampling showed a sensitivity of 80.5% (33/41 PCR positives detected; CI 66.0-89.8) and specificity of 98.6% (95% CI 94.9-99.6) compared to RT-PCR. The sensitivity with NP-sampling was 73.2% (30/41 PCR positives detected; 95% CI 58.1-84.3) and specificity was 99.3% (95% CI 96.0-100). In patients with high viral load (>7.0 log_10_ SARS-CoV2 RNA copies/swab), the sensitivity of the Ag-RDT with NMT-sampling was 100% (19/19 PCR positives detected; 95% CI 83.9-100) and 94.7% (18/19 PCR positives detected; 95% CI 76.4-99.7) with NP-sampling. In contrast, the Ag-RDT more frequently did not detect patients with lower viral load or with symptoms >7 days (Table 1), as commonly observed in studies on Ag-RDTs [7, 8].

The strengths of the study are the standardized sampling methods, two independent blinded readers and an additional semi-quantitative assessment of Ag-RDT results. The cohort was representative, judging from the comparable sensitivity observed in the recent independent validation study of STANDARD Q (sensitivity 76.6%; 95% CI 62.8-86.4) [9]. The study is limited as it was performed in a single centre. Theoretically, the previous NMT sample collection could have negatively influenced the test result of the NP sample in patients with a low viral load.

In conclusion, this study demonstrates that sensitivity of a WHO-listed SARS-CoV-2 Ag-RDT using professional nasal-sampling kit is at least equal to that of NP-sampling kit, although confidence intervals overlap. Of note, differences in the instructions for use of the test procedures could have contributed to different sensitivities. NMT-sampling can be performed with less training, reduces patient discomfort, and enables scaling of antigen testing strategies. Additional studies of patient self-sampling should be considered to further facilitate scale-up of Ag-RDT testing [6].

## Acknowledgements

Heike Rössig, Mia Wintel, Franka Kausch, Elisabeth Linzbach, Katja von dem Busche, Stephanie Padberg, Melanie Bothmann, Zümrüt Tuncer, Stefanie Lunow, Beate Zimmer, Astrid Barrera Pesek, Sabrina Pein, Nicole Buchholz, Verena Haack, Oliver Deckwart.

## Author contributions

AKL, LJK, FL and CMD designed the study and developed standard operating procedures. AKL and ON implemented the study design, enrolled patients, performed laboratory work and led the writing of the manuscript. FPM and JS coordinated and supervised the study site. CR, SB, CH, AB enrolled patients. MGe coordinated the testing facility. MGa and FT led the data analysis. TCJ and JH were responsible for PCR testing and contributed to the interpretation of the data. JAS supported the study design setup and the interpretation of the data. All authors have reviewed the manuscript.

## Data availability

All raw data and analysis code are available upon a request to the corresponding author.

